# Expected Rates of Select Adverse Events following Immunization for COVID-19 Vaccine Safety Monitoring

**DOI:** 10.1101/2021.08.31.21262919

**Authors:** Winston E. Abara, Julianne Gee, Yi Mu, Mark Deloray, Tun Ye, David K. Shay, Tom Shimabukuro

## Abstract

**Background:** Knowledge of expected rates of potential adverse events of special interest (AESI) that may occur coincidentally following COVID-19 vaccination is essential for vaccine safety surveillance and assessment. We calculated the expected rates of 21 potential AESI following COVID-19 vaccination among vaccinated persons within 1 day, 7 days, and 42 days of vaccination.

**Methods:** We used meta-analytic methods to estimate background rates of 21 medical conditions considered potential AESI and calculated expected rates of each potential AESI within 1 day, 7 days, and 42 days of vaccination.

**Results:** Background rates of three commonly monitored AESI, Guillain-Barre syndrome (GBS), myopericarditis, and all-cause deaths were 2.0 GBS cases/100,000 person-years, 1.3 myopericarditis cases/100,000 person-years, and 863.8 all-cause deaths/100,000 person-years, respectively. Based on these background rates, if 10,000,000 persons are vaccinated, we would expect 0.5, 3.7, and 22.5 GBS cases; 0.3, 2.4, and 14.3 myopericarditis cases; and 236.5, 1655.5, and 9932.8 all-cause deaths to occur in coincident temporal association (i.e., as a result of background incidence) within 1 day, 7 days, and 42 days of vaccination, respectively.

**Conclusion:** Knowledge of expected rates of potential AESI can help contextualize adverse health events associated temporally with immunization, aid in safety signal detection, guide COVID-19 vaccine public health communication, and inform benefit-risk assessments of COVID-19 vaccines.

## INTRODUCTION

The World Health Organization declared a global pandemic of coronavirus disease 2019 (COVID-19) on March 11, 2020 (1). Since COVID-19 was first detected in the United States in January 2020, more than 38 million cases and 630,000 deaths have been reported as of August 30, 2021 (2). COVID-19 vaccines are a critical tool to control and prevent COVID-19 illness (3-5). As of August 2021, one COVID-19 vaccine has received approval from the U.S. Food and Drug Administration (FDA) and two COVID-19 vaccines have received emergency use authorization. These vaccines are effective at reducing infection risk and severity of COVID-19 disease (3-5). They were developed using novel vaccine platforms. Two vaccines use mRNA encapsulated in lipid nanoparticles and the other uses a nonreplicating adenovirus vector to trigger an immune antibody response (6). Since emergency use authorization of these vaccines, large-scale COVID-19 vaccination programs have been implemented in the United States with the goal of increasing availability of and accessibility to COVID-19 vaccines to achieve community immunity in the population (7). Given the unprecedented speed of COVID-19 vaccine development, the use of novel vaccine technology, and rapid large-scale national COVID-19 vaccination, it is reasonable that people may have concerns about vaccine safety (8).

Mild to moderate local or systematic reactions after COVID-19 vaccination are common; reports of anaphylaxis and other serious adverse events such as myocarditis, thrombosis with thrombocytopenia syndrome (TTS), or Guillain-Barré syndrome (GBS) have been rare (9-11). Although a close temporal association between vaccination and an adverse event does not establish that the vaccine caused the event, such temporally associated events may lead to greater concern than more remote events. Public concern about vaccine safety can contribute to vaccine hesitancy, result in vaccine misinformation, and adversely affect vaccine uptake (12). The U.S. COVID-19 vaccine safety program comprehensively monitors the safety of all authorized COVID-19 vaccines to assess if potential adverse events are vaccine-related and exceed the expected background rate (i.e., incidence rate observed in a given population in the absence of vaccination). This monitoring is vital to ensuring safe vaccines and to promote vaccine confidence among the public.

Knowledge of the expected rates of medical conditions that would occur coincidentally (i.e., as a result of background incidence) in the U.S. population aids COVID-19 vaccine safety surveillance efforts by providing a baseline to determine if observed rates of adverse events following vaccination exceed expected rates regardless of vaccination. This information can also guide COVID-19 vaccine safety communication to the public and inform benefit-risk analyses of COVID-19 vaccines. We estimated the background rates of 21 medical conditions considered potential adverse events of special interest (AESI) following COVID-19 vaccination and calculated the expected rates of each AESI that would coincidentally occur among vaccinated persons in the general population within 1 day, 7 days, and 42 days after vaccination due to chance alone.

## METHODS

### Manuscript selection and eligibility criteria for AESI

For this analysis, we used a list of medical conditions considered potential AESI following COVID-19 vaccination from Gubernot et al. (13) (Table 1). This list was compiled by vaccine safety experts from the Centers for Disease Control and Prevention (CDC) and FDA; medical conditions considered as AESI were selected based on general or historical vaccine safety monitoring outcomes (e.g., death and GBS) and theoretical concerns related to COVID-19 disease outcomes (e.g., myopericarditis). Gubernot and colleagues reviewed the literature and presented a summary of incidence rates of these AESI that may be used in the safety surveillance and assessment of COVID-19 vaccines (13).

**Table 1.**
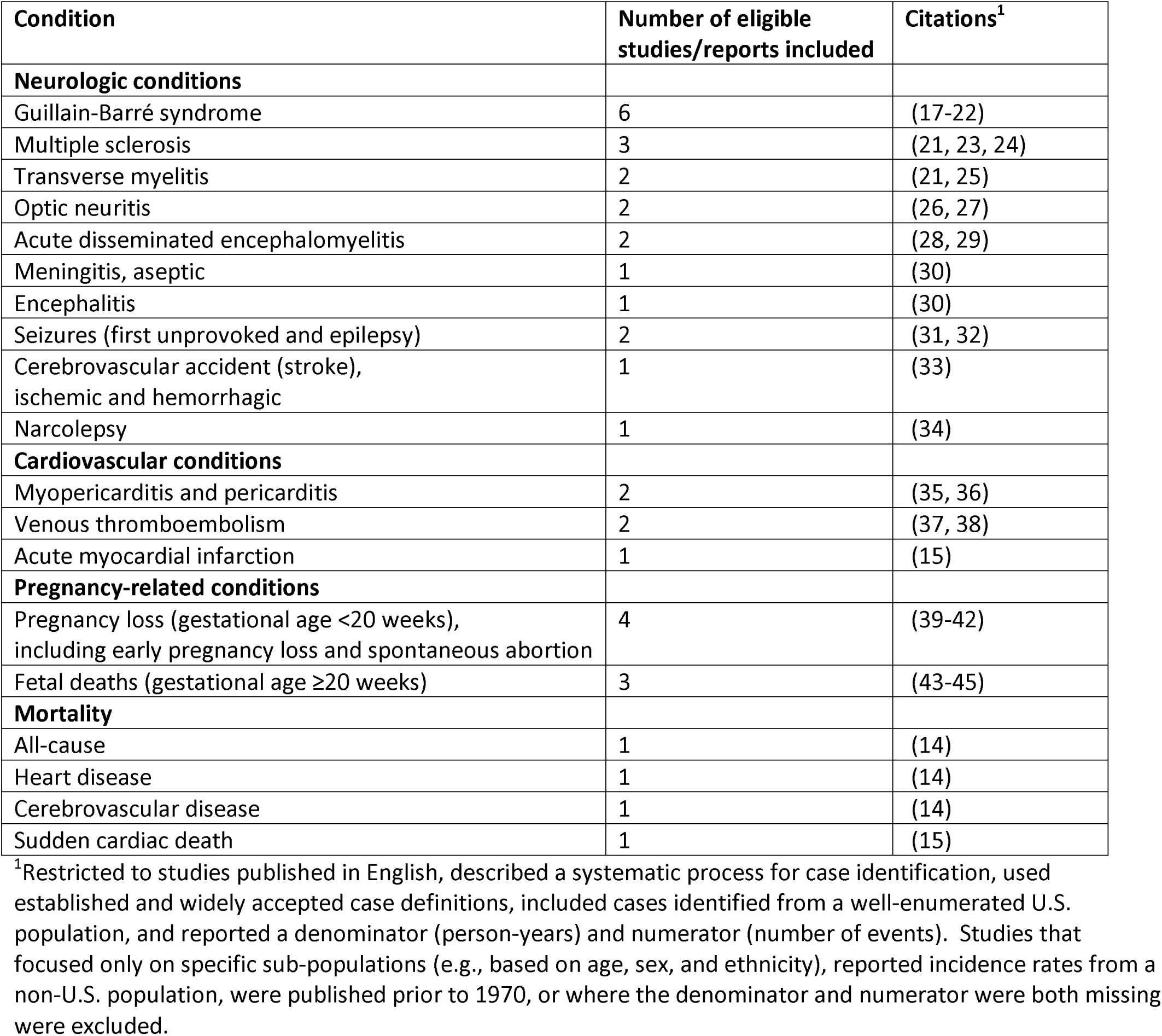
Conditions considered as potential adverse events of special interest after COVID-19 vaccination.

We reviewed the articles identified in the paper by Gubernot et al. (13). We restricted eligible studies for this analysis to those that were published in English, described a systematic process for case identification, used established and widely accepted case definitions, and included cases identified from a well-enumerated U.S. population, and reported a denominator (person-years) and numerator (number of events). If the denominator was not available in the article, we used the reported incidence rate and numerator to estimate a denominator. We excluded published articles that focused only on specific sub-populations (e.g. based on age, sex, and ethnicity), reported incidence rates from a non-U.S. population, were published prior to 1970, or where the denominator and numerator were both missing.

Rates for all-cause mortality, mortality attributed to heart disease, and mortality attributed to cerebrovascular disease were obtained from the National Vital Statistics System (NVSS) (14). The mortality rate attributed to sudden cardiac death was obtained from the American Heart Association’s Heart Diseases and Stroke Statistics 2020 update report (15). For each type of mortality event, we obtained general population rates and rates by age group and sex.

#### Analysis

We used meta-analytic methods to estimate pooled rates of potential AESI using incidence rates obtained from eligible incidence studies for each medical condition considered as an AESI after COVID-19 vaccination. We used the ‘metarate’ function in R package meta v. 4.15-1 to conduct a random effects model to estimate the pooled incidence rate and 95% confidence intervals (95% CI) for each AESI. The pooled incidence rate and 95% CI was used as the background rate for each AESI. For medical conditions where only one article met the eligibility criteria, we used the reported incidence rate and 95% CI as the background rate and 95% CI for that AESI. Because we obtained rates for all mortality events from one source, we used the reported mortality rate for each mortality event as the background rate.

### Calculating expected AESI rates

Using the derived background rates, we calculated the number of expected AESI that would occur coincidentally (i.e., not vaccine-related but as a result of background incidence) in 10,000,000 vaccinated people in the general population within 1 day, 7 days, and 42 days after vaccination with a COVID-19 vaccine. These post-vaccination time intervals were selected because the initial symptoms of an AESI can occur within hours and up to 42 days after vaccination (16). To calculate the number of expected coincident AESI, we divided the background rate by 365.25 to obtain a daily rate and by 100,000 to obtain a daily incidence rate per person. Then, we calculated the product of the daily incidence rate per person for each AESI, the number of vaccinated people (10,000,000), and the associated post-vaccination time interval (1 day, 7 days, or 42 days). For example, an AESI with a background rate of 1.87/100,000 person-years would yield a daily incidence rate per person of 0.0000000512. The product of the daily incident AESI rate per person (0.0000000512), the number of vaccinated people (10,000,000), and the post-vaccination time interval (7 days in this case) is 3.58. Thus, we would expect to observe 3.6 cases of the AESI to occur coincidentally in 10,000,000 vaccinated people in the general population within 7 days of receiving a COVID-19 vaccine, assuming a background rate of 1.87/100,000 person-years. This analysis was exempt from CDC Institutional Review Board review. The authors do not report any funding for this analysis.

## RESULTS

The calculated background rates and expected number of coincident AESI cases in 10,000,000 vaccinated persons in the general population within 1 day, 7 days, and 42 days after vaccination are summarized in the text below and in Table 2 for select AESI.

**Table 2.** Background rates and expected rates for select adverse events of special interest (number of cases/10,000,000 vaccinated people)

| **Adverse event of special interest** | **Estimated background rate (95% confidence interval)** | **Expected coincident cases within 1 day of vaccination (95% confidence interval)** | **Expected coincident cases within 7 days of vaccination (95% confidence interval)** | **Expected coincident cases within 42 days of vaccination (95% confidence interval)** |
| --- | --- | --- | --- | --- |
| Guillain-Barre syndrome | 2.0 (1.4-2.6) | 0.5 (0.4-0.7) | 3.7 (2.6-4.9) | 22.4 (15.5-29.4) |
| Multiple sclerosis | 8.9 (3.1-14.6) | 2.4 (0.9-4.0) | 17.0 (6.00-28.0) | 101.8 (35.7-168.0) |
| Transverse myelitis | 1.8 (0.0-4.3) | 0.5 (0.0-1.2) | 3.4 (0.0-8.2) | 20.1 (0.0-49.4) |
| Optic neuritis | 4.3 (3.3-5.3) | 1.2 (0.9-1.5) | 8.3 (6.3-10.2) | 49.8 (38.1-61.4) |
| Acute disseminated encephalomyelitis | 0.6 (0.5-0.7) | 0.2 (0.1-0.2) | 1.1 (0.9-1.3) | 6.6 (5.5-7.6) |
| Aseptic Meningitis | 17.8 (14.3-21.3) | 4.9 (3.9-5.8) | 34.1 (27.6-40.8) | 204.7 (164.8-244.6) |
| Encephalitis | 3.4 (2.8-4.1) | 0.9 (0.8-1.1) | 6.5 (5.3-7.8) | 39.2 (31.6-46.9) |
| 1<sup>st</sup> unprovoked seizure | 53.3 (39.0-67.6) | 14.6 (10.7-18.5) | 102.1 (74.8-129.5) | 612.6 (448.5-776.7) |
| Epilepsy (recurrent unprovoked seizures) | 38.6 (27.6-49.5) | 10.6 (7.6-13.6) | 73.9 (52.9-94.9) | 443.4 (317.7-569.2) |
| Cerebrovascular accident (stroke), ischemic and hemorrhagic | 410.2 (388.3-432.2) | 112.3 (106.3-118.3) | 786.2 (744.1-828.3) | 4717.1 (4464.5-4969.7) |
| Narcolepsy | 1.4 (0.9-1.8) | 0.4 (0.3-0.5) | 2.6 (1.8-3.5) | 15.7 (10.5-20.9) |
| Pericarditis | 7.6 (3.7-11.4) | 2.1 (1.0-3.1) | 14.6 (7.0-21.9) | 86.8 (42.2-131.4) |
| Myopericarditis | 1.3 (0.4-2.1) | 0.3 (0.1-0.6) | 2.4 (0.8-4.0) | 14.3 (5.0-23.7) |
| Venous thromboembolism | 145.1 (138.3-151.9) | 39.7 (7.9-41.6) | 278.0 (265.0-291.1) | 1668.2 (1589.9-1746.5) |
| Myocardial infarction, acute | 566.5 (565.3-567.8) | 155.1 (154.8-155.7) | 1085.8 (1083.5-1088.1) | 6514.5 (6500.3-6528.8) |
| Pregnancy loss (gestational age <20 weeks), including early pregnancy loss and spontaneous abortion | 20054.9 (17485.8-22623.9) | 5490.7 (4787.4-6194.1) | 38435.0 (33511.5-43358.6) | 230610.1 (201068.9-260151.4) |
| Fetal death (fetal death occurring after 20 weeks gestation) | 569.8 (542.6-597.1) | 156.0 (148.6-163.5) | 1092.0 (1039.8-1144.3) | 6552.2 (6238.9-6865.5) |

### Guillain-Barre Syndrome

We estimated a background rate of 2.0/100,000 person-years after a meta-analysis of six incidence studies of Guillain-Barre syndrome (17-22). Based on this background rate, we estimated that we would expect 0.5 coincident cases in 10,000,000 vaccinated persons within 1 day of vaccination; 3.7 coincident cases in 10,000,000 vaccinated persons within 7 days of vaccination; and 22.4 coincident cases in 10,000,000 vaccinated persons within 42 days of vaccination.

### Multiple sclerosis

A meta-analysis of three incidence studies of multiple sclerosis (21, 23, 24) yielded a background rate of 8.9/100,000 person-years. Assuming this background rate, we calculated that we would expect 2.4 coincident cases in 10,000,000 vaccinated persons within 1 day post-vaccination; 17.0 coincident cases in 10,000,000 vaccinated persons within 7 days post-vaccination; and 101.8 cases in 10,000,000 vaccinated persons within 42 days post-vaccination.

### Transverse myelitis

We estimated a background rate of 1.8/100,000 person-years after a meta-analysis of two incidence studies of transverse myelitis (21, 25). Based on this background rate, we would expect 0.5 coincident cases in 10,000,000 vaccinated persons within 1 day of vaccination. We would also expect 3.4 cases in 10,000,000 vaccinated persons within 7 days of vaccination and 20.1 cases in 10,000,000 vaccinated persons within 42 days of vaccination.

### Optic neuritis

A meta-analysis of two incidence studies of optic neuritis (26, 27) yielded a background rate of 4.3/100,000 person-years. Based on this background rate, we estimated that we would expect 1.2 coincident cases in 10,000,000 vaccinated persons, 8.3 coincident cases in 10,000,000 vaccinated persons, and 49.8 coincident cases in 10,000,000 vaccinated persons within 1 day, 7 days, and 42 days of vaccination, respectively.

### Acute disseminated encephalomyelitis

We estimated a background rate of 0.6/100,000 person-years for acute disseminated encephalomyelitis after a meta-analysis of two incidence studies (28, 29). Based on this background rate, we estimated that we would observe 0.2 coincident cases in 10,000,000 vaccinated persons, 1.1 coincident cases in 10,000,000 vaccinated persons, and 6.6 coincident cases in 10,000,000 vaccinated persons within 1 day, 7 days, and 42 days of vaccination, respectively.

### Aseptic meningitis

Because only one incidence rate study of aseptic meningitis was eligible for this analysis (30), we used the reported incidence rate (17.8/100,000 person-years) in this study as the background rate. Therefore, we would expect to observe 4.9 coincident cases in 10,000,000 vaccinated persons within 1 day post-vaccination; 34.1 coincident cases in 10,000,000 vaccinated persons within 7 days post-vaccination; and 204.7 coincident cases in 10,000,000 vaccinated persons within 42 days post-vaccination based on this background rate.

### Encephalitis

We used the incident rate (3.4/100,000 person-years) reported from one eligible study (30) as the background rate for encephalitis in this analysis. Assuming this background rate, we calculated that we would expect 0.9 coincident cases per 10,000,000 vaccinated persons within 1 day post-vaccination; 6.5 coincident cases per 10,000,000 vaccinated persons within 7 days post-vaccination; and 39.2 coincident cases per 10,000,000 vaccinated persons within 42 days post-vaccination.

### Seizure disorders

We categorized seizure disorders as first unprovoked seizure and epilepsy (recurrent unprovoked seizures). A meta-analysis of two incidence studies of first unprovoked seizures (31, 32) yielded a background rate of 53.3/100,000 person-years and a meta-analysis of two incidence studies of epilepsy (31, 32) yielded a background rate of 38.6/100,000 person-years. Based on the background rate of first unprovoked seizures, we expect to observe 14.6 coincident cases in 10,000,000 vaccinated persons within 1 day post-vaccination; 102.1 coincident cases in 10,000,000 vaccinated persons within 7 days post-vaccination; and 612.6 coincident cases in 10,000,000 vaccinated persons within 42 days post-vaccination.

Based on the background rate for epilepsy, we would expect to observe 10.6 coincident cases in 10,000,000 vaccinated persons within 1 day post-vaccination; 73.9 coincident cases in 10,000,000 vaccinated persons within 7 days post-vaccination; and 443.4 coincident cases in 10,000,000 vaccinated persons within 42 days post-vaccination.

### Cerebrovascular accident (ischemic and hemorrhagic stroke)

One study was eligible for this analysis and we used the reported incidence rate reported in this study (33) as the background rate for cerebrovascular accident (410.2/100,000 person-years). We would expect 112.3 coincident cases in 10,000,000 vaccinated people, 786.2 coincident cases in 10,000,000 vaccinated people, and 4717.1 coincident cases in 10,000,000 vaccinated people within 1 day, 7 days, and 42 days of vaccination.

### Narcolepsy

The incidence rate from one study (34) was used as the background rate for narcolepsy (1.4/100,000 person-years). Based on this background rate, we estimated that we would expect 0.4 coincident cases in 10,000,000 vaccinated people, 2.6 coincident cases in 10,000,000 vaccinated people, and 15.7 coincident cases in 10,000,000 vaccinated people to occur within 1 day, 7 days, and 42 days of vaccination, respectively.

### Pericarditis and myopericarditis

We estimated a background rate of 7.6/100,000 person-years for pericarditis after a meta-analysis of two incidence studies (35, 36). At this background rate, we would expect to observe 2.1 coincident cases in 10,000,000 vaccinated persons within 1 day post-vaccination; 14.6 coincident cases in 10,000,000 vaccinated persons within 7 days post-vaccination; and 86.8 coincident cases in 10,000,000 vaccinated persons within 42 days post-vaccination.

We used the incidence rate from one study (36) as the background rate for myopericarditis (1.3/100,000 person-years). At this background rate, we would expect to observe 0.3 coincident cases in 10,000,000 vaccinated persons within 1 day post-vaccination; 2.4 coincident cases in 10,000,000 vaccinated persons within 7 days post-vaccination; and 14.3 coincident cases in 10,000,000 vaccinated persons within 42 days post-vaccination.

### Venous thromboembolism

We estimated a background rate of 145.1/100,000 person-years after a meta-analysis of two incidence studies of venous thromboembolism (37, 38). Given this background rate, 39.7 coincident cases in 10,000,000 vaccinated people would be expected to occur within 1 day of vaccination; 278.0 coincident cases in 10,000,000 vaccinated people would be expected to occur within 7 days of vaccination; and 1668.2 coincident cases in 10,000,000 vaccinated people would be expected to occur within 42 days of vaccination.

### Acute myocardial infarction

We used the incident rate (566.5/100,000 person-years) reported from one eligible study (15) as the background rate for acute myocardial infarction. Based on this background rate, we estimated that we would expect 155.1 coincident cases in 10,000,000 vaccinated people, 1085.8 coincident cases in 10,000,000 vaccinated people, and 6514.5 coincident cases in 10,000,000 vaccinated people to occur within 1 day, 7 days, and 42 days of vaccination, respectively.

### Pregnancy loss (gestational age <20 weeks), including early pregnancy loss and spontaneous abortion

We estimated a background rate of 20054.9/100,000 person-years for pregnancy loss among pregnant persons (39-42). We would expect 5,490.7 cases in 10,000,000 vaccinated people within 1 day of vaccination as coincident cases based on this background rate. We would also expect 38,435.0 cases in 10,000,000 vaccinated people within 7 days of vaccination and 230,610.1 cases in 10,000,000 vaccinated people within 42 days of vaccination as coincident cases.

### Fetal death (fetal death occurring after 20 weeks gestation)

We estimated a background rate of 569.8/100,000 person-years of fetal death among pregnant persons after a meta-analysis of three studies of fetal death (43-45). We estimated that 156.0 coincident cases in 10,000,000 vaccinated people would occur within 1 day of vaccination; 1,092.0 coincident cases in 10,000,000 vaccinated people would occur within 7 days of vaccination; and 6,552.2 cases in 10,000,000 vaccinated people would occur within 42 days of vaccination assuming this background rate.

#### Mortality events

The background rates and number of expected coincident deaths attributed to all causes, heart disease, cerebrovascular disease, and sudden cardiac events overall, and by age group and gender are summarized in the text below and in Table 3.

**Table 3.** Background rates and expected rates for select mortality events.

| Mortality event | Sex | Age group (years) | Background rate (per 100,000 persons) | Expected rates within 1 day of vaccination (per 10,000,000 vaccinated persons) | Expected rates within 7 days of vaccination (per 10,000,000 vaccinated persons) | Expected rates within 42 days of vaccination (per 10,000,000 vaccinated persons) |
| --- | --- | --- | --- | --- | --- | --- |
| All-cause deaths | All | 1–4 | 24.3 | 6.7 | 12.8 | 279.4 |
|  |  | 5–14 | 13.6 | 3.7 | 26.1 | 156.4 |
|  |  | 15–24 | 74.0 | 20.3 | 141.8 | 850.9 |
|  |  | 25–34 | 132.8 | 36.4 | 254.5 | 1527.1 |
|  |  | 35–44 | 195.2 | 53.4 | 374.1 | 2244.6 |
|  |  | 45–54 | 401.5 | 109.9 | 769.5 | 4616.8 |
|  |  | 55–64 | 885.8 | 242.5 | 1697.6 | 10185.8 |
|  |  | 65–74 | 1790.9 | 490.3 | 3432.3 | 20593.5 |
|  |  | 75–84 | 4472.6 | 1224.5 | 8571.7 | 51430.3 |
|  |  | ≥85 | 13573.6 | 3716.3 | 26013.7 | 156082.5 |
|  |  | **Total** | **863.8** | **236.5** | **1655.5** | **9932.8** |
|  | Male | 1–4 | 27.3 | 7.5 | 52.3 | 313.9 |
|  |  | 5–14 | 15.6 | 4.3 | 29.9 | 179.4 |
|  |  | 15–24 | 106.1 | 29.1 | 203.3 | 1220.0 |
|  |  | 25–34 | 183.3 | 50.2 | 351.3 | 2107.8 |
|  |  | 35–44 | 249.4 | 68.3 | 477.9 | 2867.8 |
|  |  | 45–54 | 496.5 | 135.9 | 951.5 | 5709.2 |
|  |  | 55–64 | 1112.3 | 304.5 | 2131.7 | 12790.3 |
|  |  | 65–74 | 2190.2 | 599.6 | 4197.5 | 25185.1 |
|  |  | 75–84 | 5254.0 | 1438.5 | 10069.3 | 60415.6 |
|  |  | ≥85 | 14689.2 | 4021.7 | 28151.8 | 168910.7 |
|  |  | **Total** | **897.2** | **245.6** | **1719.5** | **10316.9** |
|  | Female | 1–4 | 21.1 | 5.8 | 40.4 | 242.6 |
|  |  | 5–14 | 11.4 | 3.1 | 21.9 | 131.1 |
|  |  | 15–24 | 40.4 | 11.1 | 77.4 | 464.6 |
|  |  | 25–34 | 80.8 | 22.1 | 154.9 | 929.1 |
|  |  | 35–44 | 141.4 | 38.7 | 271.0 | 1626.0 |
|  |  | 45–54 | 309.0 | 84.6 | 592.2 | 3553.2 |
|  |  | 55–64 | 674.7 | 184.7 | 1293.1 | 7758.4 |
|  |  | 65–74 | 1440.4 | 394.4 | 2760.5 | 16563.1 |
|  |  | 75–84 | 3869.1 | 1059.3 | 7415.1 | 44490.7 |
|  |  | ≥85 | 12966.5 | 3550.0 | 24850.2 | 149101.4 |
|  |  | **Total** | **831.4** | **227.6** | **1593.4** | **9560.3** |
| Deaths caused by diseases of the heart (ICD10 codes I00-I09, I11, I13, I20-I51) | All | 1-4 | 0.8 | 0.2 | 1.5 | 9.2 |
|  |  | 5-14 | 0.4 | 0.1 | 0.8 | 4.6 |
|  |  | 15-24 | 2.1 | 0.6 | 4.0 | 24.2 |
|  |  | 25-34 | 8.1 | 2.2 | 15.5 | 93.1 |
|  |  | 35-44 | 25.4 | 7.0 | 48.7 | 292.1 |
|  |  | 45-54 | 77.1 | 21.1 | 147.8 | 886.6 |
|  |  | 55-64 | 190.7 | 52.2 | 365.5 | 2192.9 |
|  |  | 65-74 | 392.9 | 107.6 | 753.0 | 4517.9 |
|  |  | 75-84 | 1028.4 | 281.6 | 1970.9 | 11825.5 |
|  |  | ≥85 | 3882.9 | 1063.1 | 7441.6 | 44649.4 |
|  |  | **Total** | **198.8** | **54.4** | **381.0** | **2286.0** |
|  | Male | 1-4 | 0.8 | 0.2 | 1.5 | 9.2 |
|  |  | 5-14 | 0.5 | 0.2 | 0.9 | 5.8 |
|  |  | 15-24 | 2.8 | 0.8 | 5.4 | 32.20 |
|  |  | 25-34 | 10.7 | 2.9 | 20.5 | 123.0 |
|  |  | 35-44 | 34.7 | 9.5 | 66.5 | 399.0 |
|  |  | 45-54 | 109.1 | 29.9 | 209.1 | 1254.5 |
|  |  | 55-64 | 273.2 | 74.8 | 523.6 | 3141.5 |
|  |  | 65-74 | 538.5 | 147.4 | 1032.0 | 6192.2 |
|  |  | 75-84 | 1306.8 | 357.8 | 2504.5 | 15026.9 |
|  |  | ≥85 | 4421.1 | 1210.4 | 8473.0 | 50838.1 |
|  |  | **Total** | **216.9** | **59.4** | **415.7** | **2494.1** |
|  | Female | 1-4 | 0.8 | 0.2 | 1.5 | 9.2 |
|  |  | 5-14 | 0.4 | 0.1 | 0.8 | 4.6 |
|  |  | 15-24 | 1.4 | 0.4 | 2.7 | 16.1 |
|  |  | 25-34 | 5.5 | 1.5 | 10.5 | 63.2 |
|  |  | 35-44 | 16.2 | 4.4 | 31.1 | 186.3 |
|  |  | 45-54 | 45.9 | 12.6 | 88.0 | 527.8 |
|  |  | 55-64 | 113.9 | 31.2 | 218.3 | 1309.7 |
|  |  | 65-74 | 265.1 | 72.6 | 508.1 | 3048.4 |
|  |  | 75-84 | 813.5 | 222.7 | 1559.1 | 9354.4 |
|  |  | ≥85 | 3589.9 | 982.9 | 6880.0 | 41280.2 |
|  |  | **Total** | **181.2** | **49.6** | **347.3** | **2083.6** |
| Deaths caused by cerebrovascular diseases (ICD10 codes I60-I69) | All | 1-4 | 0.4 | 0.1 | 0.8 | 4.6 |
|  |  | 5-14 | 0.2 | 0.1 | 0.4 | 2.3 |
|  |  | 15-24 | 0.4 | 0.1 | 0.8 | 4.6 |
|  |  | 25-34 | 1.3 | 0.4 | 2.5 | 15.0 |
|  |  | 35-44 | 4.4 | 1.2 | 8.4 | 50.6 |
| Deaths caused by cerebrovascular diseases (ICD10 codes I60-I69) |  | 45–54 | 12.3 | 3.4 | 23.6 | 141.4 |
|  |  | 55–64 | 30.3 | 8.3 | 58.1 | 348.4 |
|  |  | 65–74 | 76.4 | 20.9 | 146.4 | 878.5 |
|  |  | 75–84 | 263.1 | 72.0 | 504.2 | 3025.3 |
|  |  | ≥85 | 993.5 | 272.0 | 1904.1 | 11424.2 |
|  |  | **Total** | **44.9** | **12.3** | **86.1** | **516.3** |
|  | Male | 1–4 | 0.5 | 0.2 | 1.0 | 5.8 |
|  |  | 5–14 | 0.3 | 0.1 | 0.6 | 3.5 |
|  |  | 15–24 | 0.4 | 0.1 | 0.8 | 4.6 |
|  |  | 25–34 | 1.5 | 0.4 | 2.9 | 17.3 |
|  |  | 35–44 | 5.1 | 1.4 | 9.8 | 58.6 |
|  |  | 45–54 | 14.4 | 3.9 | 27.6 | 165.6 |
|  |  | 55–64 | 36.2 | 9.9 | 69.4 | 416.3 |
|  |  | 65–74 | 86.7 | 23.7 | 166.2 | 997.0 |
|  |  | 75–84 | 273.5 | 74.9 | 524.2 | 3145.0 |
|  |  | ≥85 | 883.3 | 241.8 | 1692.8 | 10157.1 |
|  |  | **Total** | **38.4** | **10.5** | **73.6** | **441.6** |
|  | Female | 1–4 | 0.3 | 0.1 | 0.6 | 3.5 |
|  |  | 5–14 | 0.2 | 0.1 | 0.4 | 2.3 |
|  |  | 15–24 | 0.3 | 0.1 | 0.6 | 3.5 |
|  |  | 25–34 | 1.1 | 0.3 | 2.1 | 12.7 |
|  |  | 35–44 | 3.8 | 1.0 | 7.3 | 43.7 |
|  |  | 45–54 | 10.2 | 2.8 | 19.6 | 117.3 |
|  |  | 55–64 | 24.7 | 6.8 | 47.3 | 284.0 |
|  |  | 65–74 | 67.4 | 18.5 | 129.2 | 775.0 |
|  |  | 75–84 | 255.1 | 69.8 | 488.9 | 2933.4 |
|  |  | ≥85 | 1053.4 | 288.4 | 2018.8 | 12113.0 |
|  |  | **Total** | **51.3** | **14.1** | **98.7** | **592.2** |
| Sudden cardiac death<sup>1</sup> | All | <1 | 11.2 | 3.1 | 21.5 | 128.8 |
|  |  | 1–4 | 2.2 | 0.6 | 4.2 | 25.3 |
|  |  | 5–9 | 1.2 | 0.3 | 2.3 | 13.8 |
|  |  | 10–14 | 1.2 | 0.3 | 2.3 | 13.8 |
|  |  | 15–19 | 2 | 0.6 | 3.8 | 23.0 |
|  |  | 20–24 | 3.2 | 0.9 | 6.1 | 36.8 |
|  |  | 25–29 | 5.4 | 1.6 | 10.4 | 62.1 |
|  |  | 30–34 | 8 | 2.2 | 15.3 | 92.0 |
|  |  | 35–39 | 13.3 | 3.6 | 25.6 | 152.9 |
|  |  | 40–44 | 20.9 | 5.7 | 40.1 | 240.3 |
|  |  | 45–49 | 35.6 | 9.8 | 68.2 | 409.4 |
|  |  | 50–54 | 64.8 | 17.7 | 124.2 | 745.1 |
|  |  | 55–59 | 102.3 | 28.0 | 196.1 | 1176.4 |
|  |  | 60–64 | 154.4 | 42.3 | 295.9 | 1775.4 |
|  |  | 65–69 | 220.6 | 60.4 | 422.8 | 2536.7 |
|  |  | 70–74 | 327.8 | 89.8 | 628.2 | 3769.4 |
|  |  | 75–79 | 512.4 | 140.3 | 982.0 | 5892.1 |
|  |  | 80–84 | 823.4 | 225.4 | 1578.0 | 9468.3 |
|  |  | **Total** | **97.1** | **26.6** | **186.1** | **1116.6** |

<sup>1</sup> mentioned in death certificate as contributory to death

### All-cause deaths

We used all-cause mortality rate from NVSS (14) as the background rate for all-cause deaths (863.8/100,000 persons). We estimated that we would expect 236.5 coincident all-cause deaths in 10,000,000 vaccinated people within 1 day of vaccination; 1,655.5 coincident all-cause deaths in 10,000,000 vaccinated people within 7 days of vaccination; and 9,932.8 coincident all-cause deaths in 10,000,000 vaccinated people within 42 days of vaccination. The number of expected coincident all-cause deaths in 10,000,000 vaccinated people in the general population within 1 day, 7 days, and 42 days of vaccination increased progressively by age and was consistently higher in males than in females.

### Deaths attributed to heart disease

Using heart disease mortality rate from NVSS (14) as the background rate for deaths attributed to heart disease (198.8/100,000 persons) for this analysis, we estimated that we would expect 54.4 coincident deaths attributed to heart disease among 10,000,000 vaccinated people within 1 day of vaccination; 381.0 coincident deaths attributed to heart disease among 10,000,000 vaccinated people within 7 days of vaccination; and 2,286.0 coincident deaths attributed to heart disease among 10,000,000 vaccinated people within 42 days of vaccination. The number of expected coincident deaths attributed to heart disease in 10,000,000 vaccinated people in the general population within 1 day, 7 days, and 42 days of vaccination increased progressively by age and was consistently higher in males than in females.

### Deaths attributed to cerebrovascular disease

We used cerebrovascular disease mortality rate from NVSS (14) as the background rate for deaths attributed to cerebrovascular disease (44.9/100,000 persons) for this analysis. Assuming this background rate, we expected to observe 12.3 coincident cerebrovascular disease deaths among 10,000,000 vaccinated people within 1 day of vaccination; 86.1 coincident cerebrovascular deaths among 10,000,000 vaccinated people within 7 days of vaccination; and 516.3 coincident cerebrovascular deaths among 10,000,000 vaccinated people within 42 days of vaccination. The number of expected coincident deaths attributed to cerebrovascular disease in 10,000,000 vaccinated people in the general population within 1 day, 7 days, and 42 days of vaccination increased progressively by age and was higher in females than males.

### Sudden cardiac deaths

We used the mortality rate attributed to sudden cardiac events (97.1/100,000 persons) from Virani et al. (15) as the background rate. Based on this background rate, we estimated that we would observe 26.6 coincident sudden cardiac deaths in 10,000,000 vaccinated people within 1 day of vaccination; 186.1 sudden cardiac deaths in 10,000,000 vaccinated people within 7 days of vaccination; and 1,116.6 coincident sudden cardiac deaths in 10,000,000 vaccinated people within 42 days of vaccination. The number of expected coincident deaths attributed to sudden cardiac events in 10,000,000 vaccinated people in the general population within 1 day, 7 days, and 42 days of vaccination increased progressively by age and was consistently higher in males than in females.

## DISCUSSION

Our analyses estimate expected rates of select medical conditions considered potential AESI that may occur among vaccinated persons in the general population within 1 day, 7 days, and 42 days of COVID-19 vaccination. These estimates are important for post-authorization safety monitoring of COVID-19 vaccination and for evaluating temporally associated adverse events, for framing COVID-19 vaccine safety messages more precisely, for promoting public confidence in vaccination, and for conducting benefit-risk assessment of COVID-19 vaccines.

Knowledge of expected rates of these medical conditions occurring in the general population is useful for evaluating COVID-19 vaccine safety (12, 13, 46). The observed rates of these medical conditions following vaccination can be compared to the rates that would be expected to occur coincidentally among vaccinated persons in the general population regardless of COVID-19 vaccination. This comparison can help distinguish potential vaccine-related AESI from coincidental occurrences of these adverse health events in vaccinated persons (12, 46, 47) and also complement clinical investigations by the CDC and FDA to understand the etiology of reports of potential AESI in vaccinated persons.

Expected rates of these medical outcomes in the general population can also inform public health communication about the safety of a COVID-19 vaccine (12, 13). Public confidence in vaccine safety can be negatively affected by reports that suggest a link between COVID-19 vaccination and adverse events (12). Spurious associations between vaccination and adverse health events can undermine vaccine acceptance and uptake and present challenges to achieving community immunity that is necessary for population-level protection (12). Communicating COVID-19 vaccine safety information to the public that explains that the observed rate of a potential AESI in the vaccinated population is not greater than expected can help the public understand that these events are coincident occurrences and unrelated to the vaccination (12). Framing vaccine safety communication this way may assuage vaccine safety concerns (12, 47).

Knowledge of expected rates of a potential AESI can also be useful when conducting a benefit-risk assessment of a specific COVID-19 vaccine. For example, the number of a potential AESI that is expected to occur among 10,000,000 million vaccinated persons can be compared to outcome benefits of interest such as the number of COVID-19 cases, hospitalizations, intensive care unit admissions, and deaths prevented per 10,000,000 vaccinated persons. These comparisons can assess if the population-or individual-level benefit of a specific COVID-19 vaccine outweighs the risk of a potential AESI occurring in vaccinated persons.

There are limitations to this analysis. We only calculated expected rates for a limited set of medical conditions considered as potential AESI based on the list developed by Gubernot and colleagues (13). This list does not represent the full spectrum of potential vaccine safety outcomes of concern. The generalizability of the expected rates of some potential AESI in this paper to certain subpopulations (e.g., by age, sex, and race/ethnicity) is limited because of lack of data and because rates of potential AESI can vary substantially, especially by age. We calculated expected rates of potential AESI based on background rates obtained from incidence studies, thus, there may be residual or confounding biases in the studies that we could not adequately control. The expected rates of AESI presented here are based on U.S. studies and may not be applicable to non-U.S. populations. At the time of this analysis, there were no published papers available that report the incidence estimates of TTS and myocarditis, AESI that have been associated with COVID-19 vaccination. We were therefore unable to calculate their background and expected rates.

In conclusion, knowledge of expected rates of potential AESI in vaccinated persons in the general population is vital to COVID-19 vaccine safety monitoring, distinguishing vaccine-related events from coincident events that are temporally associated with but unrelated to vaccination, informing public health COVID-19 vaccine safety communication, promoting public confidence in the vaccine, and conducting specific COVID-19 vaccine benefit-risk assessment.

## Supporting information

Supplemental checklist

## Data Availability

We conducted a meta-analysis using incidence rate data from eligible published studies cited in this paper.

## Notes

The findings and conclusions in this report are those of the authors and do not necessarily represent the official position of the Centers for Disease Control and Prevention (CDC). Authors do not declare any conflict of interests or funding sources.

### Competing Interest Statement

The authors have declared no competing interest.

### Funding Statement

There are no funding sources for this study.

### Author Declarations

This analysis was exempt from CDC Institutional Review Board review.

